# Scale-up of HIV Services for Key Population in Mozambique, 2020–2023

**DOI:** 10.1101/2024.11.25.24317615

**Authors:** Inácio Malimane, Jéssica Seleme, Gonçalves Maibaze, Lúcio Matsimbe, Irene Benech, Ana Paula Simbine, Núno Gaspar, Stélio Maposse, Josefa Mazive, Isabel Sathane, António Timbana, António Langa, Aleny Couto

## Abstract

**Introduction:** Mozambique provides targeted services to reduce HIV incidence and improve clinical outcomes among key populations (KP), including female sex workers (FSW), men who have sex with men (MSM), prisoners, and people who inject drugs (PWID). KP program data were analyzed to understand achievements since 2020 and remaining gaps.

**Methods:** We analyzed PEPFAR Monitoring, Evaluation, and Reporting routine aggregate data available for 2020–2023. KPs on antiretroviral therapy (ART) were defined as clients with ≤28 days since the last missed visit. The test positivity rate was calculated as positive tests over tests conducted. The viral load suppression (VLS) rate was calculated as KP with VLS (<1,000 copies/mL) among those tested in the past 12 months. Data were analyzed by fiscal year (October–September) and KP sub-group.

**Results:** During 2020–2023, tests conducted increased by 176% (43,834 to 121,184), positive tests increased by 33% (10,412 to 13,883) and test positivity was 17% (56,494/333,194); KP newly initiated on ART increased by 198% (3,979 to 11,875); and KP on ART increased by 370% (8,042 to 37,779). Among KP on ART in September 2023, 25,900 (69%) were FSW, 6,601 (17%) were MSM, 4,078 (11%) were prisoners, and 1,200 (3%) were PWID; VLS rate was 94% (23,224/24,723); 94% among FSW (16,451/17,551), 95% among MSM (4,195/4,419), 94% among prisoners (1,837/1,956), and 93% among PWID (741/797).

**Conclusion:** The successful scale-up of KP services in Mozambique since 2020 has increased the number of KP tested, initiated, and sustained on treatment, with high VLS rates. This indicates the positive impact of services among KP, contributing to reaching HIV epidemic control nationally.

## Introduction

As countries around the world approach HIV epidemic control, ensuring that all subpopulations are reached with non-discriminatory HIV prevention, testing, and treatment services becomes increasingly important.^1,2^ Key populations (KPs), including sex workers (SW), men who have sex with men (MSM), trans and gender-diverse people, people in prisons or other closed settings, and people who inject drugs (PWID), are particularly vulnerable to HIV and often do not have access to adequate and appropriate services.^3^ According to the Joint United Nations Program on HIV/AIDS (UNAIDS), “more than half (55%) of all new HIV infections in 2022 [globally] occurred among people from KPs and their sexual partners”.^2^

Mozambique is a high-burden country for HIV, with an estimated prevalence of 12.5% among adults aged ≥15 years, or approximately 2.4 million people living with HIV.^4^ Available data indicate that KPs in Mozambique are disproportionately affected by the HIV epidemic.^2^

Starting in 2020, the Mozambique Ministry of Health (MISAU), with support from the U.S. President’s Emergency Plan for AIDS Relief (PEPFAR), increased the number of sites offering KP-friendly services, trained health providers, hired KP peer navigators, updated KP guidelines and registries, and expanded care and prevention services (e.g., provision of pre-exposure prophylaxis (PrEP), condoms, and safe lubricant) at each entry point in health facilities to ensure that all KPs are offered tailored services.

This evaluation aims to analyze KP program data to describe how Mozambique has scaled up the provision of KP-friendly services since 2020 and identify remaining gaps.

## Methods

We conducted a retrospective analysis of PEPFAR Monitoring, Evaluation, and Reporting (MER) data for KP programming reported for fiscal years 2020–2023 in all sites supported by PEPFAR.

Individuals were classified into one of the following based on their reported behaviors and gender identity: female sex workers (FSW), MSM, transgender women, and PWID. People in prisons were defined by their current incarceration status. Test positivity was calculated as the number of positive tests divided by the number of tests conducted. Linkage to antiretroviral therapy (ART) was assessed using a proxy indicator consisting of the number of newly identified KPs with HIV infection who initiated ART divided by the number of positive tests. KPs newly initiated on ART were those with an HIV-positive test who started ART and did not have a previous history of ART. KPs on ART were defined as KP clients with an ART pharmacy pick-up ≤28 days after their last clinic visit. Retention was defined as the proportion of KP clients who remained on ART among those who initiated treatment at least 12 months prior to the evaluation period. The proxy indicator for viral load coverage was calculated as the number of KPs with a viral load test over the number of KPs on ART 12 months before the evaluation period. Viral load suppression (VLS) was defined as a viral load test result of <1,000 copies/mL, and the VLS rate was calculated as the number of KPs with VLS among those with viral load tests conducted in the past 12 months. Data were extracted from DATIM and analyzed by fiscal year (October–September), KP sub-group, age group (<25 years, ≥25 years), and province using Excel.

This activity was reviewed by CDC, deemed not research, and conducted consistent with applicable federal law and CDC policy. <u>^1^</u>

## Results

From 2020 to 2023, annual tests conducted among KPs increased by 169% (42,484 to 114,480) (Table 1). The number of positive tests increased by 51% (9,522 to 14,414), and the test positivity rate decreased from 22.4% (9,522/42,484) to 12.6% (14,414/114,480) (Table). Among the 114,480 tests conducted in fiscal year 2023, the test positivity rates by KP sub-group were 18.6% (9,235/49,689) among FSW, 10.8% (2,565/23,782) among MSM, 5.6% (1,819/32,384) among prisoners, 8.9% (657/7,352) among PWID, and 10.8% (138/1,273) among transgender women. The median test positivity rate among KPs by province was 12.6% (range = 6.6% [1,462/22,072] in Maputo Province to 18.2% [420/2,308] in Tete Province).

**Table 1:** Mozambique Key Population (KP) Program Indicators and Percentage Change, Fiscal Years 2020 and 2023.

| Indicators | Fiscal Year |  | % Change |
| --- | --- | --- | --- |
|  | 2020 | 2023 |  |
| Tests conducted | 42,484 | 114,480 | 169.4% |
| Positive tests | 9,522 | 14,414 | 51.3% |
| Test positivity rate | 22.4% | 12.6% | -43.7% |
| % Proxy linkage | 42% | 82.4% | 95.2% |
| Newly initiated on antiretroviral therapy | 3,979 | 11,875 | 198.4% |
| On treatment | 8,042 | 37,779 | 369.7% |
| Viral load tests conducted | 2,029 | 23,224 | 1044.6% |
| Viral load tests w/ viral load suppression (<1,000 copies/mL) | 2,362 | 24,723 | 946.7% |
| % Proxy viral load testing coverage | 92.4% | 74.7% | -18.4% |
| % Viral load suppression | 85.9% | 93.9% | 9.3% |
| % Retention at 12 Months | No data | 86.6% | - |

Overall, 34,541 KP clients were newly initiated on ART from 2020 to 2023 (FSW = 23,501 [68%]; MSM = 5,849 [17%]; prisoners = 3,784 [11%]; PWID = 1,404 [4%]), and the annual number newly initiated increased by 198% (3,979 to 11,875). In fiscal year 2023, the proxy linkage rate was 82.4% (11,875/14,414) among FSW, 85.1% (7,876/9,253) among MSM, 86.9% (2,231/2,565) among prisoners, 65.2% (1,186/1,819) among PWID, and 88.1% (579/657) among transgender women. The median proxy linkage rate by province for KPs was 82% (range = 50.5% [594/1,174] in Inhambane Province to 115.5% [4,251/3,680] in Nampula Province).

The number of KP clients on ART increased by 370% (8,042 to 37,779) from 2020 to 2023. Among KP on ART in September 2023, 25,900 (69%) were FSW, 6,601 (17%) were MSM, 4,078 (11%) were prisoners, and 1,200 (3%) were PWID. By September 2023, the 12-month retention rate was 83.9% (2,464/2,938); the retention rate was 85.1% among FSW (1,744/2,048), 84.7% among MSM (502/593), 74.3% among PWID (110/148), and 72.4% among prisoners (108/149). The median 12-month KP client ART retention rate by province was 87% (range = 75% [2,616/3,509] in Maputo City Province to 96% [3,383/3,527] in Manica Province).

In September 2020, the overall proxy viral load testing coverage was 92.4% (2,362/2,557). In September 2023, this rate decreased to 74.7% (24,723/33,091); it was 77.6% (17,551/22,612) among FSW, 76.1% (4,419/5,804) among MSM, 53.2% (1,956/3,675) among prisoners, and 79.7% (797/1,000) among PWID. The median proxy viral load testing coverage among KP clients by province was 75% (range = 56% [1,795/3,194] in Sofala Province to 89% [659/739] in Niassa Province).

In September 2020, the overall viral load suppression rate among KPs was 85.9% (2,029/2,362). By September 2023, this rate increased to 93.9% (23,224/24,723); it was 93.7% (16,451/ 17,551) among FSW, 94.9% (4,195/4,419) among MSM, 93.9% (1,837/1,956) among prisoners, and 92.9% (741/797). The median viral load suppression rate by province was 94% (range = 89% [1,051/1,176] in Inhambane Province to 97% [1,045/ 1,080] in Gaza Province).

## Discussion

### Program Growth and Service Expansion

Mozambique has demonstrated significant growth in the number of KPs reached with HIV testing, care, and treatment services between fiscal years 2020 and 2023. During this period, the program diagnosed over 35,000 KPs living with HIV and initiated nearly 30,000 individuals on ART. This expansion, while commendable, revealed disparities across geographical regions and KP subpopulations, necessitating a closer examination of the factors contributing to these differences.

### HIV Testing

The number of KPs tested for HIV increased substantially during the study period, reflecting the program’s efforts to expand testing coverage. However, this increase was accompanied by a decline in the positive yield, which dropped from 22.4% in 2020 to 12.6% in 2023. This decline may be attributed to several factors, including the saturation of testing in higher-prevalence areas and the inclusion of lower-risk individuals in the testing pool. To counteract this trend and improve case finding, the program may need to implement more tailored testing strategies, particularly in areas and subpopulations where HIV prevalence remains high, balancing testing volume with testing efficiency as indicated by test positivity rates.

### Linkage to Care

Linkage to care is a critical step in the HIV treatment cascade, and while the program achieved a notable increase in linkage rates overall, 95%, significant disparities persist across KP subgroups. For example, PWID and prisoners exhibited lower linkage rates compared to FSW and MSM. The high proxy linkage rate of over 100% observed in Nampula could be due to testing or identification of KPs who are living with HIV outside of the PEPFAR-supported health information systems. For example, KPs may be identified by community partners at the community level or through registers at general health facilities and then referred to treatment services at PEPFAR-supported sites offering KP-specific care and treatment services, thus increasing the proxy linkage rate through inflated numbers of newly initiated individuals compared to number of positive tests.

### ART Retention

Retention in ART programs is essential for achieving sustained viral suppression and preventing HIV transmission. As of September 2023, the overall retention rate for KPs at 12 months was 87%. However, this rate was notably lower among PWID (74%) and prisoners (72%), indicating a need for more robust follow-up and support mechanisms. These subgroups may face unique challenges, such as stigma, mobility, and lack of social support, which can hinder their ability to remain engaged in care. Addressing these challenges through targeted interventions, such as the allocation of dedicated peer navigators and enhanced post-release follow-up for prisoners, could improve retention rates. Additionally, implementing harm reduction programs, particularly opioid substitution therapy (OST) for PWID, is crucial for enhancing retention by providing comprehensive support that addresses their specific needs.

### Viral Load Coverage and Suppression

While improved over the years, viral load testing coverage remains an area of concern, particularly given the decrease from 92% in 2020 to 75% in 2023. One contributing factor to this decline is the increased capture of key populations in our registries due to the introduction of new HIV registration tools, which expanded the follow-up cohort and brought similar challenges to those faced by the general population in providing comprehensive services, such as viral load testing. Additionally, the expansion of differentiated service delivery models during the same period contributed to inadequate follow-up regarding the testing schedule, leading to delays in viral load sample collection. These factors underscore the need for ongoing efforts to optimize viral load testing coverage and ensure timely and appropriate monitoring for all KPs. Prisoners had the lowest proxy viral load coverage at 53%, which affected understanding of viral load suppression outcomes among this KP. Despite these challenges, the program substantially increased the number of KPs with viral load suppression, demonstrating the effectiveness of ART when KPs are retained in care. It is vital to consider targeted efforts to address barriers to viral load testing to further enhance viral load coverage and suppression, particularly among prisoners and provinces with low coverage, such as Sofala (56%). These efforts could greatly benefit from increasing access to testing facilities, improving the coordination between correctional and health services, and providing targeted education to encourage testing among eligible KPs.

### Limitations

This study has several limitations that should be considered when interpreting the findings. Program data are subject to data quality issues, although ongoing data quality assurance activities aim to mitigate these concerns. Additionally, the identification of KPs relied on self-reported risk behaviors and gender identity, which may have led to misclassification due to stigma and fear of discrimination. Moreover, the use of proxy indicators may not fully capture the actual performance. Despite these limitations, the study provides valuable insights into the progress and challenges of KP service delivery in Mozambique.

## Conclusion

The scale-up of KP-friendly HIV services in Mozambique has resulted in substantial improvements along the entire HIV cascade, from case finding to treatment initiation and viral load suppression. Over three years, the number of KP clients on ART increased by 370%, reaching nearly 40,000 KPs nationwide.

Addressing ongoing challenges in service delivery can sustain these gains and continue progressing toward epidemic control. Enhanced linkage to care, particularly among subgroups like PWID and prisoners, where linkage rates remain lower, and improved retention and viral load testing coverage—especially in regions and subpopulations with the most significant gaps can close service gaps.

Ongoing investment in tailored interventions that address the unique needs of each KP subgroup helps ensure sustained service delivery to KPs. Increased access to KP-friendly services, enhanced follow-up mechanisms, particularly for those transitioning out of high-risk environments like prisons, and resource allocation for continued support and capacity-building among healthcare providers are evidence-based approaches that may help Mozambique strengthen its HIV response and achieve the UNAIDS 95-95-95 goals among KPs.

## Data Availability

All data produced are available from PEPFAR Monitoring, Evaluation, and Reporting program data.

https://data.pepfar.gov/datasets#PDD

## Authors’ contributions

Inácio Malimane, Irene Benech, Gonçalves Maibaze and Lúcio Matsimbe conceived the study, drafted the manuscript, and conducted the analysis; Ana Simbine, António Timbana, Josefa Mazive, and Stelio Maposse drafted the methods; and Nuno Gaspar, Jessica Seleme, António Langa, and Aleny Couto drafted the discussion and conclusion. All authors contributed to a critical review of the manuscript.

## Acknowledgments

We would like to acknowledge the support of PEPFAR Mozambique, the Mozambican Ministry of Health, the National AIDS Council, and the National Health Institute.

## Footnotes

1 See e.g., 45 C.F.R. part 46.102(I)(2), 21 C.F.R. part 56; 42 U.S.C. 241(d); 5 U.S.C. 552a; 44 U.S.C. 3501 et seq.

